# Emotion regulation and cognitive function as mediating factors for the association between lifetime abuse and risky behaviors in women of color

**DOI:** 10.1101/2023.01.07.22283182

**Authors:** Karina Villalba, Lisa H. Domenico, Robert L. Cook, Julia O’Connor, Kyndester Michael-Samaroo, Maria Jose Del Pino Espejo, Pilar Martin, Jessy G. Dévieux

## Abstract

**Background:** Lifetime abuse (i.e., childhood abuse, intimate partner violence) has been linked to risky behavioral outcomes (i.e., alcohol use, risky sex). Women who experience lifetime abuse have poor emotion regulation and may have problems with executive functioning, which could help explain the relationship between lifetime abuse and risky behaviors. However, research on executive functioning and emotion regulation as mediators of this relationship has been limited. In the present study, we examined this association. We hypothesized that lifetime abuse would be related to greater difficulty in emotion regulation and executive function, which would also be associated with greater alcohol use and risky sex.

**Methods:** This cross-sectional study included 150 women with a history of lifetime abuse who were assessed for alcohol use using the AUDIT Score; emotion regulation was measured using the Difficulties with Emotion Regulation Scale (DERS); risky sex was measured using question: “in the last 90 days, how many people did you have anal or vaginal sex without using a condom? Executive function was assessed using the NIH Toolbox.

**Results:** The mediation model followed the self-regulation theory, which proposes executive function as the higher-order cognitive process. Results showed that executive function deficit and poor emotion regulation significantly mediated the relationship between lifetime abuse and alcohol use severity (indirect effect = .097, SE .031, 95% CI = .035 to .158).

**Conclusion:** Our findings suggest executive function and emotion regulation as a potential mechanism for alcohol use severity in women who experienced lifetime abuse (i.e., childhood abuse and intimate partner violence).

## 1. Introduction

One of the long-term adverse implications of childhood abuse is revictimization in adulthood, with higher rates among women who experienced childhood abuse than among women who did not experience childhood abuse (1, 2). There is increasing support from the research literature to view lifetime abuse as a cumulative effect of different types of abuse (e.g., emotional, physical, and sexual) from one or various perpetrators across the developmental lifespan [e.g., childhood, adolescence, adulthood; ((3-6)].

Lifetime abuse is associated with multiple negative outcomes, including alcohol use disorders (AUD) and risky sex. Women who experience lifetime abuse are more likely to report high-risk partners (e.g., intravenous drug users, HIV-positive individuals). They tend to engage in risky sexual behaviors, including inconsistent condom use, promiscuity, and early sexual initiation, increasing their risk for HIV infection (7-9). There is also growing evidence suggesting that women who experienced lifetime abuse can be less sexually assertive. This behavior can result in less condom insistence and a greater likelihood of unprotected sex (10). Additionally, women who experienced lifetime abuse are twice as likely to suffer from AUD compared to women who experienced either childhood abuse or intimate partner violence alone (11-13). These women are more likely to begin drinking at a younger age and report more problems related to alcohol use (14-16).

In addition, women who experienced lifetime abuse have long-term difficulties with emotion regulation strategies and engage in emotionally avoidant coping behaviors ((17-21)). Furthermore, Furthermore, women in violent relationships are usually subjected to emotional invalidation by their partners, which in turn can lead to developing more frequent and long-lasting periods of inability to control their emotions and use alcohol as a negative coping behavior (22, 23). Women who experienced lifetime abuse are also at a higher risk of developing cognitive problems, including executive dysfunction. Developmental neuroscience suggests that adverse environments resulting from abuse and/or violence may expose children to chronic stress, which can disrupt brain architecture and impair the development of executive function, leading to cognitive problems in adulthood (24).

The Self-regulation Model proposes a causal order that includes executive function as a higher-order cognitive process that contributes to emotion regulation (25). This higher-order model (i.e., executive function promoting emotion regulation) provides a framework for understanding how individuals interpret, respond, and adjust to negative experiences. Self-regulation failure is associated with negative emotional states, resulting in executive function deficit and poor emotional regulation, ultimately leading to maladaptive coping behaviors (26). Executive function is described as a higher-order cognitive process that includes planning, cognitive flexibility, social cognition, emotion regulation, and behavioral competencies such as attitudes and behaviors (Miyake et al., 2000). Executive function can be described as either hot or cold. Cold executive functions are associated with planning, working memory, behavioral monitoring, and inhibition. Whereas hot executive functions are conceptualized as social cognition and emotions (Chung et al., 2011). There has been increased interest in recent years in the role of hot executive functioning as the control center of emotional expression. This is important because these processes are crucial for an individual’s social interaction and social-emotional development (Banducci et al., 2014). Emotion regulation can be intrinsic or extrinsic; intrinsic refers to one regulating their own emotions (i.e., self-regulatory processes); extrinsic refers to an individual regulating someone else’s emotions (e.g., a parent regulating the emotions of an infant by soothing her (27). Developmental psychology studies have shown that one of the roles of a parent is to teach a child emotion regulation strategies; however, children growing up in abusive environments may not be able to develop an intrinsic regulation mechanism because of the lack of support from their parents (28, 29). In fact, research shows that adult survivors of childhood abuse exhibit poorer intrinsic emotion regulation skills than their non-abused counterparts (30). Thus, poor emotion regulation denotes a rigid and maladaptive use of emotion regulation strategies and the inability to choose the most appropriate strategy for achieving goals (31).

Research indicates well-established relationships between childhood abuse, intimate partner violence, poor emotion regulation, and risk behaviors (alcohol abuse and risky sex). (13, 25, 32). However, the present study aimed to provide additional evidence for an association between cognitive function, emotion regulation, and risky sex and propose that deficit in cognitive function and poor emotion regulation mediates the relationship between lifetime abuse and risk behaviors (alcohol abuse and risky sex). Furthermore, the present study adds to the literature in multiple ways. First, we address lifetime abuse conceptualized as emotional, physical, or sexual revictimization across two periods (childhood and adulthood). This is important as much of the prior research has focused on a single type of abuse and/or time period. Second, we anticipate that poor emotion regulation and deficit in cognitive function will be indirectly associated with lifetime abuse and risk behaviors. Previous research shows that women who experienced abuse may be at increased risk for relying on maladaptive strategies for self-regulating negative emotions or coping with negative experiences. As such, we expect that lifetime abuse would be significantly associated with difficulty in emotion regulation. Therefore, we propose to evaluate executive function and emotion regulation as mediating pathways in the relationship between lifetime abuse and risk behavioral outcomes. We hypothesized that poor emotion regulation and a greater deficit in executive function would mediate the association between lifetime abuse, alcohol use severity, and risky sex.

## 2. Methods

### 2.1. Participants and Procedure

This was a cross-sectional study with 150 women of color (African American, Hispanic) reporting a history of childhood abuse and intimate partner violence between 2018 and 2019.

The sample consisted of women of color (Hispanic and African American) at risk for HIV recruited from four community-based organizations located in a densely populated, multicultural, urban area in South Florida. The recruitment was in-person, using flyers and presentations. To be eligible, participants had to be between 18 and 50 years old; have a history of childhood abuse and/or intimate partner violence; have at least one episode of a sexual encounter in the last three months; a history of alcohol use in the last three months; fluent in spoken English and/or Spanish; and be able to provide informed consent. This study was approved by the Institutional Review Board of Florida International University, and all women provided written informed consent.

### 2.2 Measures

The survey was available in both English and Spanish. The survey instruments that were not validated in Spanish were translated into Spanish and then back-translated and reviewed by a Cultural Linguistic Group to ensure cultural appropriateness and relevance. The survey instrument measured the following domains used within the current analysis.

#### Demographics

Self-reported sociodemographic information included age, race and ethnicity, employment status, and education.

### 2.3 Predictors

#### Intimate partner violence

The World Health Organization (WHO) Self-reported Questionnaire (33) was used to measure intimate partner violence. This is a culturally adapted questionnaire from a multi-country study conducted by the WHO. The 20-item questionnaire asked participants about the type of abuse (emotional, physical, sexual) and the timing of the abuse (last 12 months, lifetime). Responses to these questions were ‘Yes (= 1)’ and ‘No (= 0)’. All questions were added to compute a composite score that ranged from 0 to 13 for IPV, with higher scores indicating higher abuse.

#### Childhood Abuse

The Childhood Trauma Questionnaire measured childhood emotional, physical, and sexual abuse and neglect (Fink et al., 1995). Participants were asked about their experiences with types of abuse during their childhood. All questions utilized a five-point Likert scale with responses ranging from “never true” to “very often,” with a total of four subscales measuring each type of abuse with a maximum possible score of 25, with higher scores reflecting higher childhood abuse.

#### Lifetime abuse

For this study, we developed a continuous additive weighted composite score that included variables from childhood abuse and intimate partner violence.

### 2.4 Mediators

#### Difficulties with Emotion Regulation

This construct was measured using the Difficulties with Emotion Regulation (DERS) scale (Gratz & Roemer, 2003), which evaluates emotion regulation difficulties in six areas, including nonacceptance of emotional responses, difficulties engaging in goal-directed behaviors, impulse-control difficulties, lack of emotional awareness, limited access to emotion regulation strategies, and lack of emotional clarity. Scores ranged between 36 to 180, with prior research suggesting the clinical range on DERS total scores between 80 -127. Higher scores indicate greater difficulties with emotion regulation.

#### Executive function

was measured using the NIH Toolbox in English and Spanish using normative data from English-and Spanish-speaking populations (34). Executive control was measured using the Flanker Inhibitory Control and Attention and the Dimensional Change Card Sort test. We pooled the Flanker Test, which measures the allocation of an individual’s limited capacities to deal with environmental stimulation, and the Dimensional Change Card Sort Test, which measures the capacity to plan, organize and monitor the executive of behaviors that are strategically directed, in a goal-oriented manner as predictors of executive functioning. Higher scores indicated higher levels of cognitive functioning. A standard score at or near 100 indicates cognitive ability that is average compared with others nationally. Standard scores around 115 suggest above-average cognitive ability, while scores around 130 suggest superior ability (in the top 2 percent nationally, based on Toolbox normative data). Conversely, a standard score of around 85 suggests below-average cognitive ability, and a score in the range of 70 or below (bottom 2 percent) suggest very low cognitive functioning.

### 2.5 Outcomes

#### Risky sex

To measure sexual risk, we used a continuous variable using the following question: “in the last 90 days, how many people did you have anal or vaginal sex with without using a condom?”

#### Alcohol use severity

The Alcohol Use Disorders Identification Test (AUDIT) is a 10-item survey that measures alcohol consumption, dependence symptoms, and personal and social harm reflective of drinking over the past 30 days (Saunders et al., 1993). It covers the areas of alcohol consumption, drinking behavior, and alcohol-related problems. Responses are scored from 0 to 4, with a maximum possible score of 40, with higher scores indicating problematic drinking. A score of 8 or more is associated with harmful or hazardous drinking. For this study, hazardous alcohol use was a continuous variable defined as hazardous alcohol use with a score ≥ 8.

## 3. Data Analysis

The data analytic plan included three steps. First, descriptive statistics were computed; second, correlations were estimated for all variables in the mediation models; and third, to measure the alcohol and risky sex mediating models, we measured the direct and indirect associations between lifetime abuse, executive function, emotion regulation, alcohol use, and risky sex.

Descriptive data were analyzed using SPSS 26.0 (IMB, Chicago, IL) and were checked for normality, outliers, and missing values. Descriptive statistics (mean, standard deviations, skewness, kurtosis) were calculated to describe sample characteristics and tested for normality assumptions for all continuous variables. Correlation analyses were conducted for all continuous variables. Bivariate analysis of demographic and key variables was conducted utilizing chi-square tests. Demographic and other continuous variables significantly associated at the bivariate level with alcohol use severity/risky sex were included in subsequent models as covariates.

We performed two serial mediation models – Model 1, alcohol use severity, and Model 2, risky sex. These models used the serial mediation model to predict the relationship between executive function deficit and poor emotion regulation as mediating variables in the pathway between lifetime abuse and risk behaviors. The direct and indirect effects of lifetime abuse on alcohol use severity and risky sex via the mediating mechanism of executive function and emotion regulation were assessed with multiple ordinary least squares (OLS) regressions through the SPSS macro PROCESS version 3.203 (Hayes & Preacher, 2014). This method simultaneously estimates the direct association of X on Y (c’-path), the direct association of X on M (a-path), the direct association of M on Y (b-path), and the indirect association of X (lifetime abuse) on Y (alcohol use severity or risky sex) via M (poor emotion regulation and executive function deficit). The indirect effect (i.e., mediation) was tested using 10,000 resampling bias-corrected bootstrap confidence intervals (95% CI). Indirect effect models were run separately for alcohol use severity and risky sex. Lifetime abuse was entered as the X (independent) variable.

Executive function deficit and poor emotion regulation were entered as serial mediating variables into each model [alcohol use severity and risky sex (i.e., number of partners in the last 90 days). Alcohol use severity and risky sex were entered into the models individually as the Y (dependent) variable. Age and level of education were entered into all models as control variables. We chose the OLS regression as a preferred method as it offers the least Type I and Type II errors (Preacher & Hayes, 2004), and it has greater power to detect mediational effects than similar approaches (MacKinnon et al., 2002).

## 4. Results

### 4.1 Descriptive Analysis

The mean age was 42 (SD = 11.0) years old (see Table 1). Forty-seven percent were Black, with 54% of the sample self-identified as Hispanic. The mean alcohol use among the women was 10.4 (SD = 8.6), indicating alcohol use severity. The mean number of sexual partners without using a condom in the last 90 days was 2 (SD = .97). The mean score for emotion regulation was 84.2 (SD = 24.5), with higher scores indicating difficulties in emotion regulation; similarly, the executive function score of 72 (SD = 12.5) indicated lower executive functioning compared to the normative data which may indicate difficulties in general functioning.

**Table 1.**
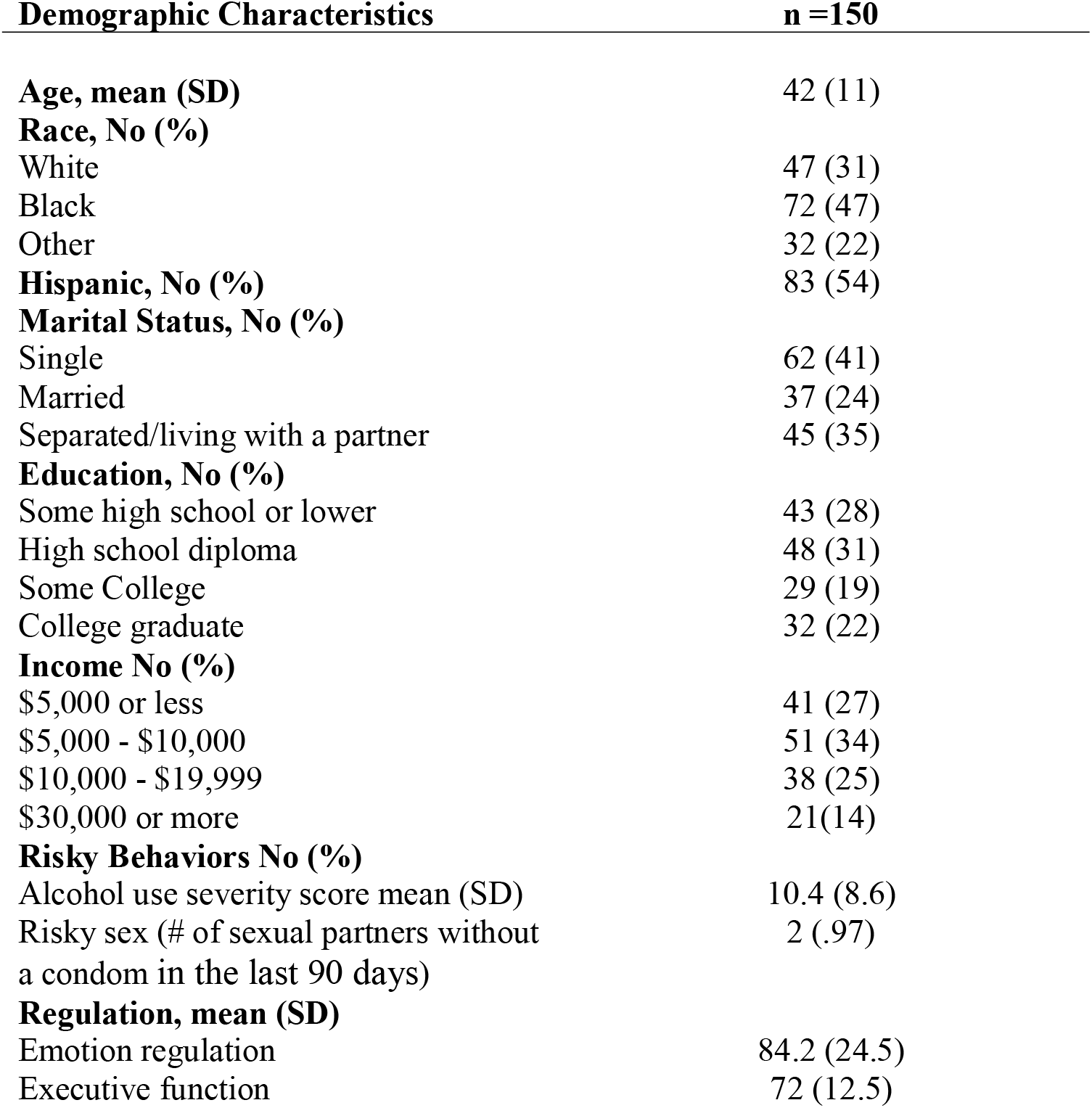
Sociodemographic Characteristics Among Women of Color with Lifetime Abuse

| **Demographic Characteristics** | **n =150** |
| --- | --- |
| **Age, mean (SD)** | 42 (11) |
| **Race, No (%)** |  |
| White | 47 (31) |
| Black | 72 (47) |
| Other | 32 (22) |
| **Hispanic, No (%)** | 83 (54) |
| **Marital Status, No (%)** |  |
| Single | 62 (41) |
| Married | 37 (24) |
| Separated/living with a partner | 45 (35) |
| **Education, No (%)** |  |
| Some high school or lower | 43 (28) |
| High school diploma | 48 (31) |
| Some College | 29 (19) |
| College graduate | 32 (22) |
| **Income No (%)** |  |
| \$5,000 or less | 41 (27) |
| \$5,000 - \$10,000 | 51 (34) |
| \$10,000 - \$19,999 | 38 (25) |
| \$30,000 or more | 21(14) |
| **Risky Behaviors No (%)** |  |
| Alcohol use severity score mean (SD) | 10.4 (8.6) |
| Risky sex (# of sexual partners without a condom in the last 90 days) | 2 (.97) |
| **Regulation, mean (SD)** |  |
| Emotion regulation | 84.2 (24.5) |
| Executive function | 72 (12.5) |

### 4.2 Correlational Analysis

We found a positive correlation between lifetime abuse and poor emotion regulation (.284, *p* < .001). Risky sex was positively correlated with poor emotion regulation (.324, *p* < .001) and alcohol use severity (.183 *p* < .05; See Table 2).

**Table 2.** Study variables correlation analysis

|  | **1** | **2** | **3** | **4** | **5** | **6** |
| --- | --- | --- | --- | --- | --- | --- |
| **(1) Age** | . |  |  |  |  |  |
| **(2) Risky Sex** | **-.196\*** | . |  |  |  |  |
| **(3) Lifetime abuse** | -.068 | .144 | . |  |  |  |
| **(4) Poor emotion regulation** | .069 | **.324\*\*** | **.284\*\*** | . |  |  |
| **(5) Executive function deficit** | .116 | -.101 | .138 | -.188 | . |  |
| **(6) Alcohol use severity** | **.183\*** | **.250\*\*** | .129 | **.505\*\*** | **.325\*\*** | . |

### 4.3 Mediation Results

We tested two serial mediation models with differing outcome variables. Model 1 - alcohol use severity; Model 2 – risky sex. The mediators were ordered between the predictor and the outcome. The order of the mediators followed the self-regulation theory; we first included executive function as the higher-order cognitive process, followed by emotion regulation (26).

Since this is a cross-sectional study, we cannot imply causality, but results may pave the way for future longitudinal mediation analysis measuring key study constructs.

### 4.4 Alcohol Use Severity

For Model 1, the total effect model was statistically significant; the mediators, executive function deficit, and poor emotion regulation were indirectly associated with lifetime abuse and alcohol use severity. See Figure 1 which illustrates the results of the mediation model analysis and Table 3 for the direct and indirect results. This was a fully mediated pathway. The indirect effect of executive function deficit via poor emotion regulation on the association between lifetime abuse and alcohol use severity was significant, with the total effect size accounting for 10% for a deficit in executive functioning and emotion regulation (indirect effect = .097, SE .031, 95% CI = .035 to .158).

**Table 3:**
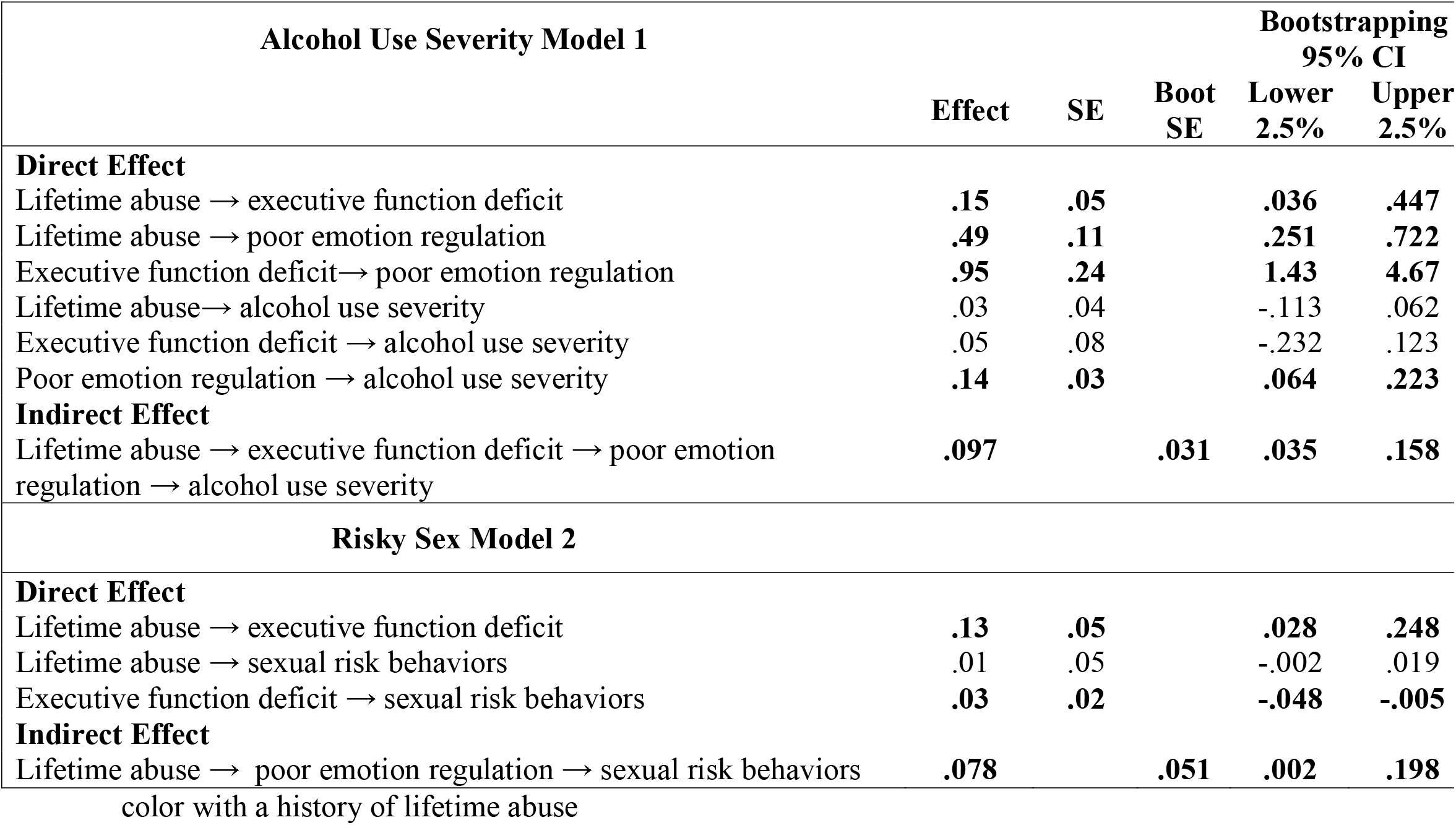
Direct and Indirect analysis for alcohol use severity and risky sex among women of

**Figure 1:**
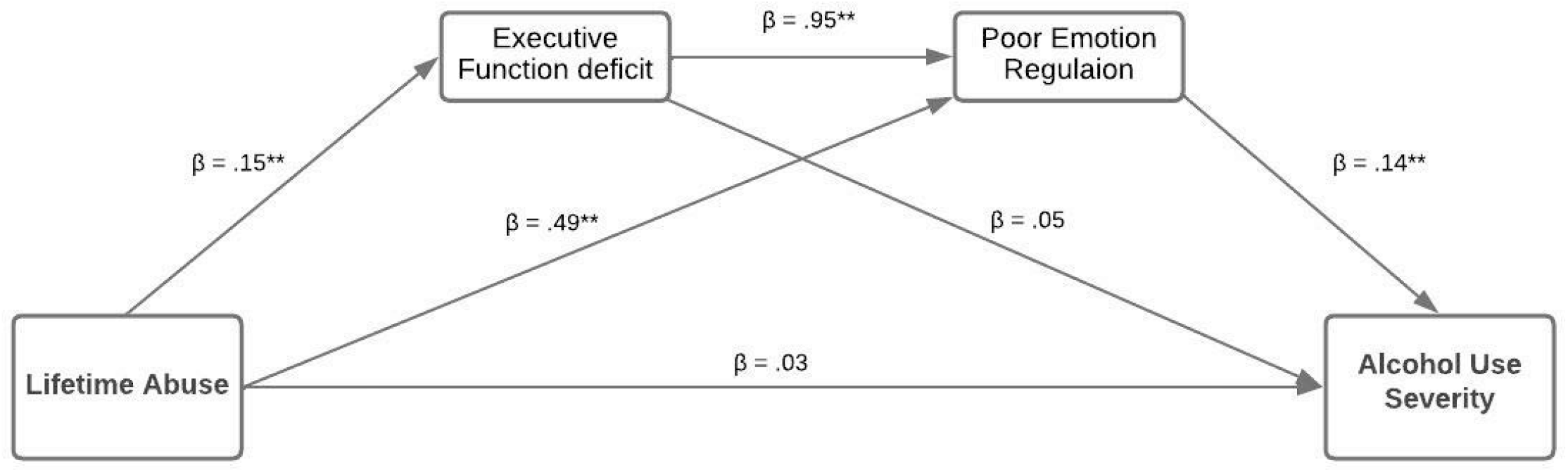
Mediation analysis between lifetime abuse, executive function deficit, poor emotion regulation, and alcohol use severity among women of color among women of color

### 4.5 Risky Sex Model

For the serial multiple mediation risky sex (Model 2), we measured the mediating effect of executive function deficit and poor emotion regulation between lifetime abuse and risky sex. The results of the model were partially significant. Only a deficit in executive function was indirectly associated with lifetime abuse and risky sex. See Figure 2 which illustrates the results of the mediation model analysis and Table 3 for the direct and indirect results. This was a fully mediated pathway. The indirect effect via deficit in executive function on the association between lifetime abuse and risky sex was significant, with the total effect size accounting for 8% (indirect effect = .078% SE = .051 95% CI = .002 to .198).

**Figure 2:**
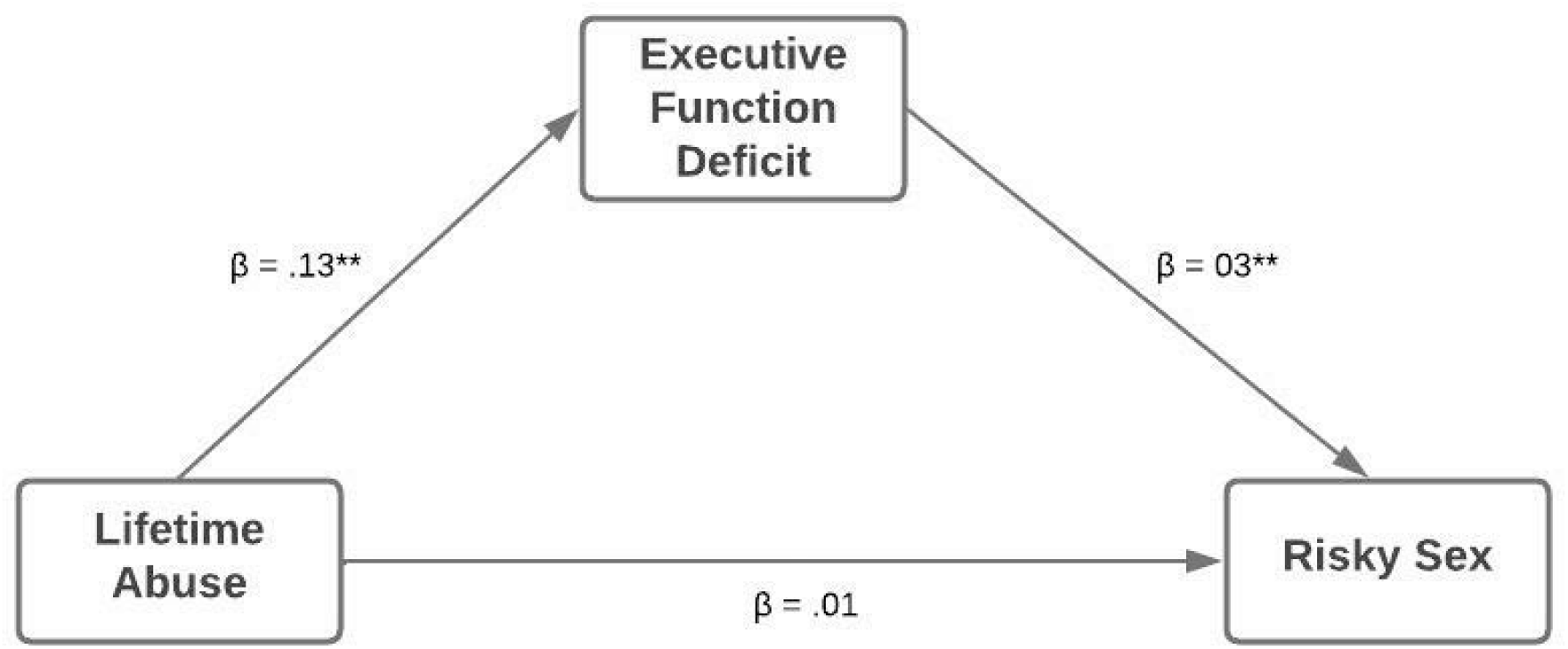
Mediation analysis between lifetime abuse, executive function deficit, and risky sex among women of color

## 5. Discussion

To gain further insight into the pathway between lifetime abuse and risky behavioral outcomes, we evaluated the mediating role of executive function and emotion regulation using a serial mediation model. This study has the potential to significantly add to the understanding of the mechanisms that contribute to increased alcohol use and risky sex in women of color with a history of lifetime abuse. For these models, we followed the self-regulation theory, which focuses on a higher-order relationship between emotions and cognition. Consistent with our hypothesis, our results support previous findings in the literature regarding alcohol use (35-37); for women with lifetime abuse, reported poor emotion regulation was associated with greater alcohol use severity. Our study is also in line with a recent study demonstrating that exposure to childhood abuse was associated with a deficit in executive function and poor emotion regulation, which predicted future alcohol use behaviors in adolescents (38). Thus, our findings suggest that the pathway between lifetime abuse and alcohol use severity may occur through a cognitive-emotional mechanism. Results from this study are also consistent with the self-regulation theory, which proposes a higher-order cognitive process in connection with emotion regulation to produce an integrated behavioral response (Mueller & Peterson, 2012). Emerging research shows that growing up in an abusive environment contributes to a deficit in executive functioning and poor emotion regulation. These types of environments can affect the organization and integration of neural circuits involved in emotion and cognitive functions, which can impair self-regulation and contribute to a host of maladaptive outcomes in adulthood (24, 28-30). Thus, the pathway to emotionally avoidant coping behaviors (e.g., alcohol use severity) may be through a deficit in executive functioning and heightened emotional reactivity in women with a history of lifetime abuse.

Although we did not identify specific executive function processes or emotion regulation facets, this study demonstrated that the mechanism for alcohol use severity in women with lifetime abuse could be explained via the cognitive-emotional model. Research focusing on specific processes of executive function and emotion regulation may present valuable information on the mechanism for self-regulation among women with lifetime abuse.

We also tested the mediating role of executive function deficit and poor emotion regulation in the relationship between lifetime abuse and risky sex. We hypothesized that a deficit in executive function and poor emotion regulation would mediate the relationship between lifetime abuse and risky sex; however, our results showed that executive function deficit was the only significant mediator. Although more research needs to be done to understand the effect of emotion regulation on risky sex, the results showed that lower executive functioning is significantly associated with increased risky sex. Neuroscience research has shown that alterations in the frontoparietal executive control network in adults with a history of abuse have implications for decision-making (Zhao et al., 2021). Moreover, impairments in the executive control network lead to a shift from flexible, goal-directed behavior to inflexible stimulus-response, making self-regulation more difficult (39). Another factor that may have the potential to influence risky sex is cognitive appraisals. For example, anticipated negative partner reaction has predicted women’s decreased sexual assertiveness and condom self-efficacy (40). Our study was not designed to determine factors that may have contributed to this behavior. However, this is the first known study to analyze these relationships; thus, a longitudinal study focusing on specific executive function and emotion regulation processes for risky sex may be warranted.

### 5.1 Limitations and Future Directions

Findings from this investigation should be interpreted in light of its limitations. First, although we used validated self-report measures commonly used in the field, in-person interviews are subject to recall and social desirability bias. Second, we did not use specific processes to measure executive function and emotion regulation; instead, we used the cumulative assessment for these two variables. As mentioned earlier, disentangling the effects of executive function and emotion regulation systems will offer a better understanding of how these relationships may predict such outcomes. Third the cross-sectional design precludes conclusions about causal inferences and mediating effects. However, the independent variable of interest in the present study was collected retrospectively based on past exposure to abuse, while the mediators and outcomes were collected based on the present results. Therefore, the foundation for temporal ordering is present and adds support for the current design (41, 42). Fourth, these variables may not be the only mediators in the models. It could be that there are other mediators which may not be in the model or even in the dataset. Nevertheless, we frame our findings in terms of indirect effects and suggest that the present study findings provide theoretical contributions and lay the foundation for further longitudinal mediation analysis of the key study constructs. While we certainly acknowledge these various limitations, this work represents the early stages of a novel approach to understanding an understudied population. Finally, the sample size was small. Future studies with a larger sample size may be able to test whether emotion regulation mediates risky sex and confirm the cognitive-emotional model.

This is the first known study analyzing the indirect effects of executive function deficit and poor emotion regulation on lifetime abuse and risk behaviors which provides a framework for understanding current work but also highlights how much remains to be clarified in future research. Although there is emergent research on the ability to regulate emotions, cognition, and behavior in response to internal and external demands in various populations, only one known prior study measured executive function as the pathway between early life adversity and performance (43). It will be important to replicate our findings with a greater focus on the specific processes of executive function and emotion regulation related to self-regulation. It would also be important to include an interactive approach to the data analysis to clarify how different components of executive function and emotion regulation support the response to internal and/or external factors.

### 5.2 Conclusions

This study measured the mediating role of executive function and emotion regulation in the relationship between lifetime abuse and alcohol use severity and risky sex. Our findings suggest executive function and emotion regulation as a potential mechanism for alcohol use severity in women who experienced lifetime abuse (i.e., childhood abuse and intimate partner violence). Conversely, the indirect pathway for risky sex was only significant through executive function. We cannot determine the reason for this relationship, but we offered a few theories and proposed that a longitudinal study may provide further clarification. In sum, we showed that women exposed to lifetime abuse may have a deficit in their self-regulation mechanism through the cognitive-emotional pathway, thus relying on maladaptive strategies such as alcohol use and risky sex for regulating their negative emotions.

## Data Availability

All relevant data are within the manuscript and its Supporting Information files.

https://www.ucf.edu/

